# Predicting Olfactory Loss In Chronic Rhinosinusitis Using Machine Learning

**DOI:** 10.1101/2020.10.12.20210500

**Authors:** Vijay R. Ramakrishnan, Jaron Arbet, Jess C. Mace, Krithika Suresh, Stephanie Shintani Smith, Zachary M. Soler, Timothy L. Smith

## Abstract

**Objective:** Compare machine learning (ML) based predictive analytics methods to traditional logistic regression in classification of olfactory dysfunction in chronic rhinosinusitis (CRS-OD), and identify predictors within a large multi-institutional cohort of refractory CRS patients.

**Methods:** Adult CRS patients enrolled in a prospective, multi-institutional, observational cohort study were assessed for baseline CRS-OD using a smell identification test (SIT) or brief SIT (bSIT). Four different ML methods were compared to traditional logistic regression for classification of CRS normosmics versus CRS-OD.

**Results:** Data were collected for 611 study participants who met inclusion criteria between April 2011 and July 2015. 34% of enrolled patients demonstrated olfactory loss on psychophysical testing. Differences between CRS normosmics and those with smell loss included objective disease measures (CT and endoscopy scores), age, sex, prior surgeries, socioeconomic status, steroid use, polyp presence, asthma, and aspirin sensitivity. Most ML methods performed favorably in terms of predictive ability. Top predictors include factors previously reported in the literature, as well as several socioeconomic factors.

**Conclusion:** Olfactory dysfunction is a variable phenomenon in CRS patients. ML methods perform well compared to traditional logistic regression in classification of normosmia versus smell loss in CRS, and are able to include numerous risk factors into prediction models. Several actionable features were identified as risk factors for CRS-OD. These results suggest that ML methods may be useful for current understanding and future study of hyposmia secondary to sinonasal disease, the most common cause of persistent olfactory loss in the general population.

## INTRODUCTION

Olfactory loss is common, affecting up to 25% of the adult population, with chronic rhinosinusitis (CRS) being a major cause of persistent olfactory loss [1]. Smell loss is a hallmark of CRS, particularly in those with nasal polyps, and is one of the cardinal diagnostic symptoms for the disease [2-4]. Curiously, this symptom does not affect all CRS patients. Just as CRS is a heterogeneous disease [5], previous literature has estimated a 20-80% prevalence of olfactory loss in CRS cohorts, based on the sensitivity of olfactory assessment used and the proportion of enrolled subjects with nasal polyps (CRSwNP). A recent study using well-defined CRS diagnostic criteria and Sniffin’ Sticks objective testing showed a 44% prevalence of smell loss in CRSsNP subjects and 58% in CRSwNP [6]. Risk factors associated with CRS-OD identified in this study included disease severity score on computed tomography (CT) and the presence of comorbid conditions such as diabetes, allergy, and asthma. Adding a healthy control group for comparison, Schlosser et al. noted CRS-OD to affect CRSwNP patients more than CRSsNP, and identified age as an additional risk factor [7]. Given that most existing studies consist of single-institution cohorts, with subjects undergoing variable treatment regimens, and different assessments of smell function, a systematic review and meta-analysis was performed by Kohli et al. [8] to clarify the presence of OD in CRS and identify consistent risk factors. The authors included 47 studies in the analysis, and identified age, CT score, and presence of nasal polyps as the three most important risk factors for CRS-OD in the published literature. Similarly, olfactory response to CRS treatment is highly variable, with resolution of olfactory disturbance after standard therapies occurring in approximately 40% of refractory CRS patients [9].

Olfactory dysfunction in CRS is potentially complex and the interplay among possible mechanisms is poorly understood. The simple model of conductive loss due to impaired odorant transport to the olfactory cleft in CRS may explain a portion of the disease for some patients. Local and systemic inflammation likely contribute to varying degrees in particular individuals, with underlying factors such as age and presence of comorbid diabetes influencing the capacity for olfactory sensory neuron regeneration. Given the intricate and multifactorial nature of CRS-OD, compounded by inherent challenges of human cohort studies, it is no surprise that putative predictors and their relative importance have been inconsistently characterized across studies. Existing single-institution studies carry a significant risk of sampling bias and their use of univariate regression models are insufficient to identify predictor variables for CRS-OD. These analyses cannot account for all possible predictor variables or interactions among variables, and therefore, may offer limited insights. Additional attempts at categorizing outcomes with multivariable regression approaches that were not developed for predictive performance, or assessed as such, cannot be relied upon [10].

Traditional statistical approaches can model how system variables relate to one another and generate corresponding mathematical metrics of statistical significance. However, novel data analytics approaches including machine learning (ML) methods have shown improved classification accuracy and can be utilized to predict outcomes, rather than focusing on individual components within a system. ML approaches are particularly useful when there is a role for including complex interactions that may otherwise be ignored or dismissed as noise when using traditional statistics [11], such as in the case of CRS-OD.

Understanding the heterogeneity of CRS-OD and interpreting multi-dimensional risk factors for outcomes prediction is foundational for future clinical care and research. The objective of this study was to test different ML-based predictive analytics approaches against traditional regression modeling for classification accuracy of OD in CRS patients. A secondary goal was to explore top predictors in high-performing models to identify common predictor variables of importance that may help explain the heterogeneity of OD found among CRS patients.

## MATERIALS and METHODS

### Study Population

Adult patients (≥18 years old) were recruited into a prospective, multi-center, observational cohort study, for which multiple reports have been previously published [12-15]. All study patients were diagnosed with medically refractory CRS as defined by the American Academy of Otolaryngology-Head and Neck Surgery 2007 guidelines [4]. Performance sites consisted of tertiary care sinus centers located within academic hospital systems in North America including: Oregon Health & Science University (OHSU; Portland, OR; eIRB#7198), Stanford University (Palo Alto, CA; IRB#4947), and the Medical University of South Carolina (Charleston, SC; IRB#12409), with central study coordination at OHSU. The Institutional Review Board (IRB) at each performance site governed all study protocols, informed consent documentation, and data safety monitoring, in accordance with the Declaration of Helsinki for Medical Research Involving Human Subjects.

At original study enrollment, participants were screened for demographics, social and medical history (**Table 1**). The diagnoses of asthma, allergy, and aspirin-exacerbated respiratory disease (AERD) were made by evaluation of the medical record, presence of classic symptoms and self-report of prior testing. Subjects with comorbid cystic fibrosis, ciliary dyskinesia, or autoimmune disease were included in the analysis. Patient reported outcome measures (PROMs) were collected during study enrollment, but were not used in the predictive analytics given the overlap between particular symptom ratings and objective smell loss [22-25]. Given the exploratory, retrospective nature of this study, all available subjects were included and sample size calculations were not conducted.

**Table 1.**
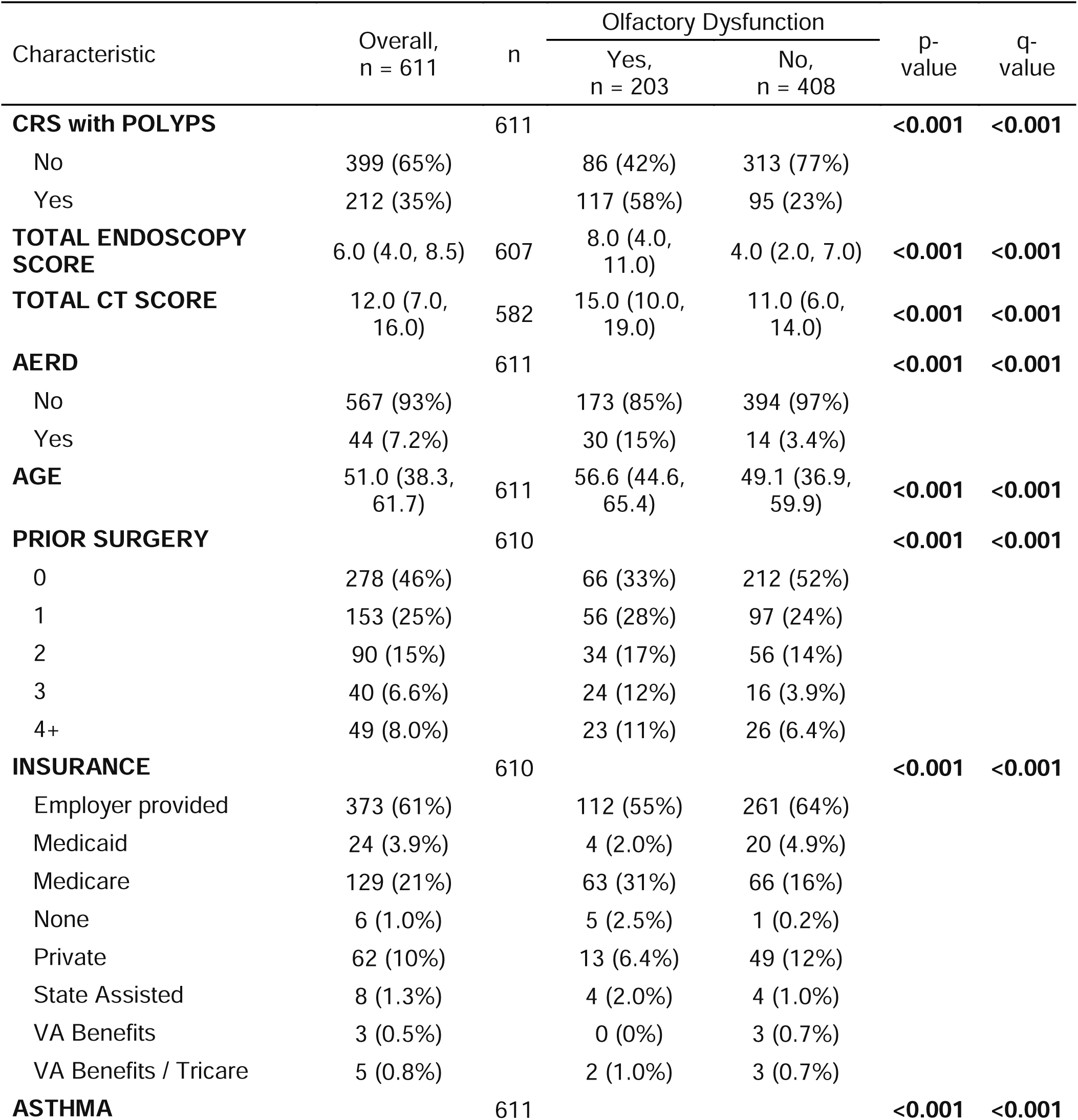
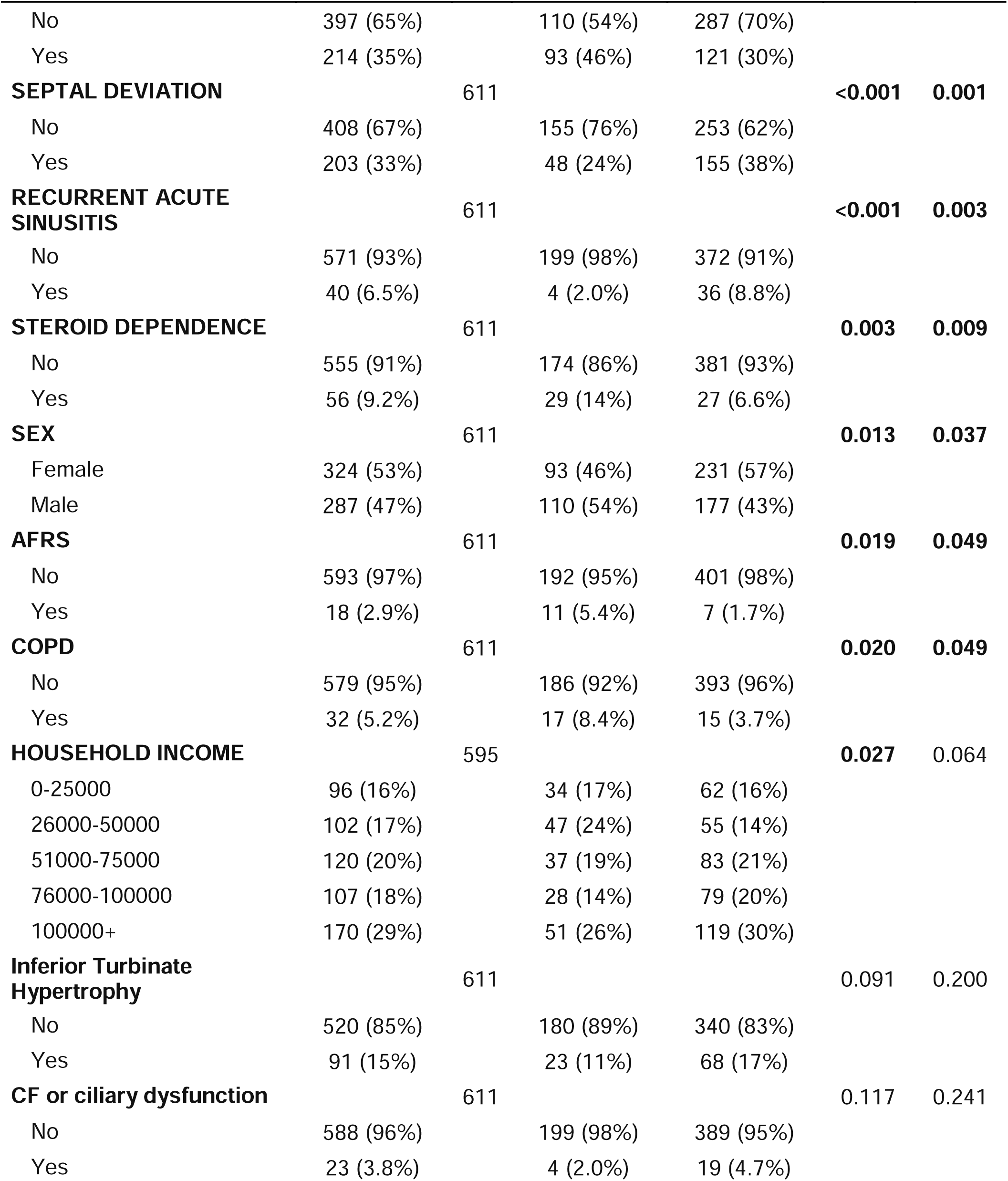
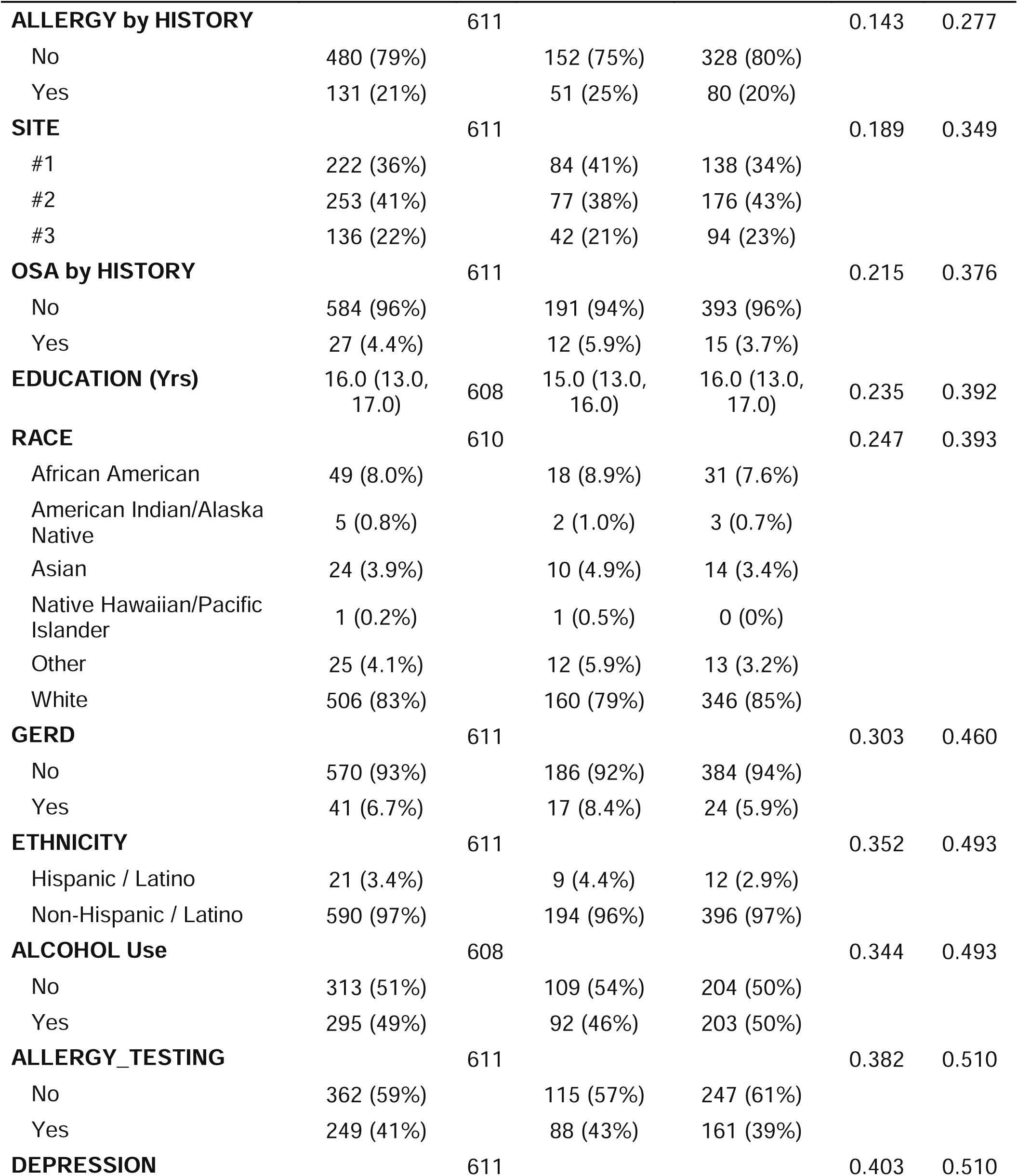
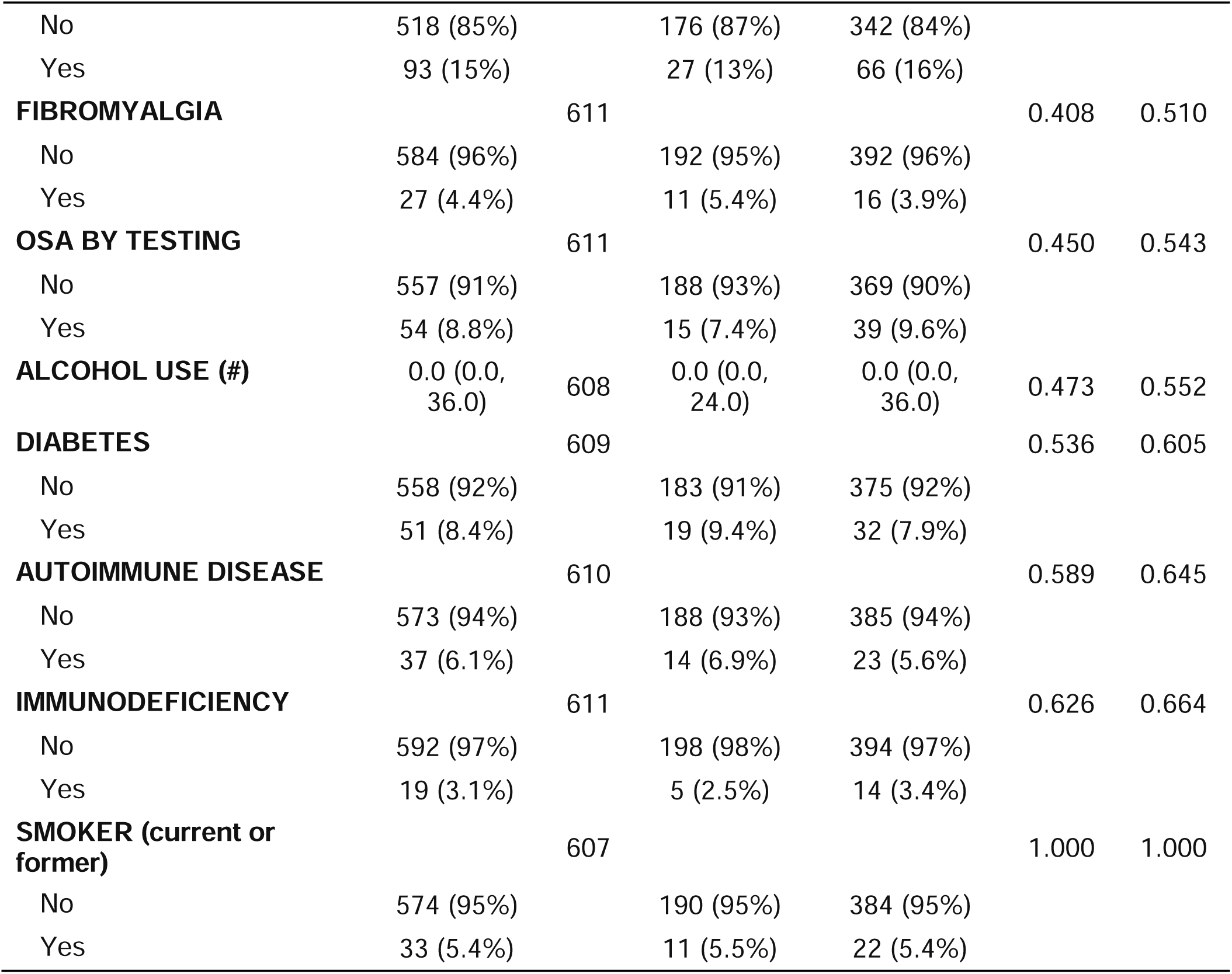
Study population. Collected demographics and clinical metadata used as inputs for classification models are shown here for the overall cohort, and those with and without olfactory dysfunction. Count (%) and Fisher’s exact test were used for categorical variables, while median (25^th^ quantile, 75^th^ quantile) and Wilcoxon rank sum test were used for numeric variables. Data sorted from smallest to largest q-value (multiple testing adjusted p-value).

### Objective Measures of Disease Severity

Diagnostic measures of disease severity were collected as standard of care, and were also used for investigational purposes. These well-established objective measures have been recommended for clinical study [16], and include Lund-Kennedy nasal endoscopy scoring [17] and high-resolution CT imaging graded using the Lund-Mackay scoring system [18].

The main outcome of interest in the current study is olfactory dysfunction as measured by psychophysical testing. Olfactory function was initially assessed using the brief Smell Identification Test (bSIT) in the years 2011-2013, and subsequently using the 40-item Smell Identification Test (SIT) in the years 2013-2015 (Sensonics Inc., Haddon Heights, NJ). Higher SIT total scores reflect a better sense of smell, and categorical ratings of “smell loss” (anosmia + hyposmia) and “normosmia” were assigned based on established scales [19-21].

### Data Management and Statistical Analyses

Protected health information was previously removed, and study data were safeguarded using a unique study identification number assignment for each participant. Study data were securely transferred from a HIPAA compliant, relational database (Access, Microsoft Corp, Redmond, WA.) to the University of Colorado Department of Otolaryngology-Head and Neck Surgery password protected research server, according to specifications of a Data Use Agreement between OHSU and the University of Colorado.

Study data were evaluated descriptively using count (%) for categorical variables and median and interquartile range (IQR; 25^th^ quantile, 75^th^ quantile) for numeric variables. Comparing variables between normosmics and those with smell loss, Fisher’s exact test was used for categorical variables and Wilcoxon rank sum test was used for numeric variables. Both the original p-values and Benjamini-Hochberg multiple testing adjusted p-values (“q-values”) are reported [26]. Statistical comparisons with q-values < 0.05 were regarded as statistically significant. All statistical and data analyses were completed using the R software program [27]. Machine learning models were compared to simpler logistic regression model with backwards step-wise variable selection using Akaike information criteria (AIC). The step-wise variable selection was re-performed during each iteration of the cross-validation procedure (details later), in order to prevent biased estimates of classification accuracy [28].

### Machine Learning Approaches to Classification

Four different machine learning predictive analytics models were applied, in order to compare classification accuracy to logistic regression, and code was made publicly available at https://github.com/vijayramakrishnan/chemsenses-CRS-olfactory-prediction.git. All classifiers were fit using the caret R package [29], which provides a unified framework for tuning and evaluating classification accuracy. When including a categorical predictor in a model, K-1 dummy variables were used, where K is the total number of categories. *Random forests (RF)* was chosen as it allows for non-linear and interaction effects, and can handle a large number of potential predictors [30]. Although the original random forest algorithm is biased towards favoring features that have more possible cut-points (e.g. categorical variables with more categories), importantly, we used a special type of random forests for unbiased feature selection: an ensemble of “conditional inference trees” with permutation-based variable importance scores [31-33]. Similar to traditional logistic regression, least absolute shrinkage and selection operator (*LASSO*) assumes only linear and additive effects (without interaction) where regression coefficients are shrunk toward zero to perform variable selection [34,35]. In this model, unimportant variables are given coefficients equal to zero, effectively removing them from the model. Along these lines, multivariate adaptive regression splines (*MARS*) was also applied in a similar fashion to traditional logistic regression, but it uses stepwise methods for variable selection and allows for nonlinear and interaction effects [36]. Lastly, the *Support Vector Machine* (*SVM*) approach was applied with a radial basis kernel, which allows for non-linear and interaction effects [37]. Although many machine learning methods exist, these four methods were selected since RF and SVM with a Gaussian/radial basis kernel have been shown to perform well across a wide variety of empirical applications, while LASSO and MARS were chosen as simpler, easier to interpret methods [38-40]. Given that the primary goal is to compare how well different models classify a binary olfactory dysfunction outcome, unsupervised machine learning methods (which do not use an outcome) were not considered appropriate for this study.

Repeated 10-fold cross validation (repeated 5 times [41]) was used to tune and internally validate the classification accuracy of each model. The following measures of accuracy were considered: area under receiver operating characteristic curve (AUC), sensitivity (proportion of subjects with olfactory dysfunction that were correctly classified), and specificity (proportion of subjects without olfactory dysfunction that were correctly classified). To deal with the fact that the two classes are unbalanced (dysosmia vs normosmia), under-sampling was used within the cross-validation procedure, as implemented in the caret R package [29,42]. Over-sampling was also considered, but produced nearly equal classification accuracy in terms of AUC (results not shown). The method of “surrogate splits” [33] was used to handle missing values in RF. The missForest R package [43] was used to impute missing values for all other models, while excluding the outcome in order to prevent over-fitting. Cross-validated AUC was compared between models using a corrected resampled t-test [44], which corrects for the overlap that occurs during the resampling process, and the Benjamini-Hochberg method was used to adjust for multiple testing.

To identify the important predictor variables included in each classification algorithm, permutation-based variable importance scores were used to rank variables from most to least important in RF, while the absolute value of the standardized regression coefficient was used for LASSO. The IML R package [45] was used to calculate variable importance scores for SVM using a permutation method. All variable importance scores were scaled from 0 to 100, where the most important variable for the model has a score of 100. Of note, for a categorical variable with more than two categories, RF and LASSO calculate separate variable importance scores for each category, whereas the approach used for SVM only calculates an overall score.

## RESULTS

### Final Cohort Characteristics

A total of 611 study participants met all inclusion criteria and were prospectively enrolled between April 2011 and July 2015. Demographics and comorbid disease characteristics, examined by olfactory classification, are described in **Table 1**. The overall median age for the final cohort was 51 years (IQR= 38.3, 61.7) years with slight female preponderance (53% female). Approximately one-third of the cohort exhibited nasal polyps (35%).

Study participants were stratified by olfactory testing category. Statistically significant differences (q<0.05) were observed between the normosmic and hyposmic cohorts in terms of objective disease measures (CT and endoscopy scores), age, sex, previous surgery, socioeconomic status, corticosteroid dependence, asthma/COPD, and AERD. Sinonasal conditions including nasal polyps, allergic fungal rhinosinusitis, and deviated septum were also significant factors.

### Smell loss vs Normosmia Classification Accuracy

**Figure 1** illustrates a comparison of the different models based on AUC, sensitivity, and specificity. A logistic regression model for classification of baseline olfactory dysfunction demonstrated acceptable accuracy in the context of CRS (AUC 0.707±0.07). Three of the four machine learning models performed favorably compared with the traditional logistic regression model. The SVM approach was the most accurate, with a relatively good classification accuracy (AUC 0.754±0.05) and much higher sensitivity than other the methods. LASSO and Random Forest also outperformed logistic regression, whereas MARS had the lowest AUC. The corrected resampled t-test with Benjamini-Hochberg adjustment (**Figure 1c**) indicated that SVM had a significantly higher mean AUC than MARS (q=0.02), but was not significantly different than the Random Forest, LASSO, or logistic regression. LASSO had a significantly higher mean AUC than MARS (q=0.012) and logistic regression (q=0.012).

**Figure 1.**
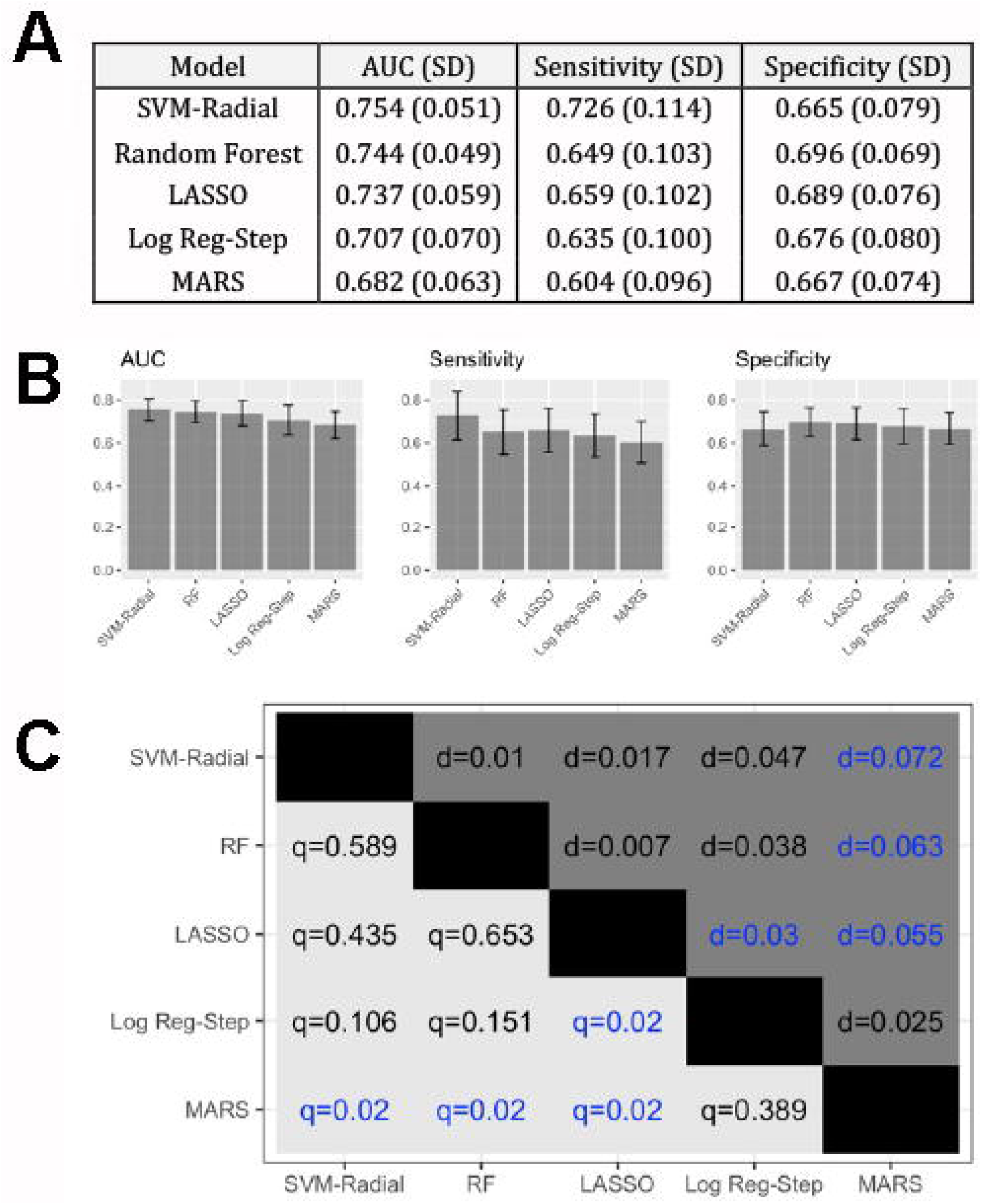
Comparing AUC between methods. **A and B) Classification accuracy for the different models.** The sample mean and standard deviation of the performance metrics is reported across all 50 cross-validation resampling estimates. **C) Statistical comparison of methods**. Upper triangle depicts the mean difference in AUC (d) between the model on the y-axis and the model on the x-axis. Lower triangle depicts the q-value (multiple testing adjusted p-value) for the corrected resampled t-test. Blue values indicated statistically significant differences in mean AUC between the corresponding models on the x-axis and y-axis. *AUC = area under receiver operating characteristic curve. SVM-Radial = support vector machine with a radial basis kernel; RF = random forest; Log Reg-Step = logistic regression with step-wise variable selection*.

### Machine Learning Predictions: Top variables

ML algorithms are adept at recognizing patterns within complex data, but the lack of *a priori* framework results in the clinical challenge of understanding *why* the algorithm made its decisions. The ML models carried some different relative importances of predictor variables. SVM, the model with the largest AUC, considered many variables to be influential, with 20 variables having variable importance scores >25 (**Fig 2**). In Random Forest and LASSO models, fewer variables were important and the most weight was placed on the presence of nasal polyps and the nasal endoscopy severity score (**Fig 3 and Suppl Fig 1**).

**Figure 2.**
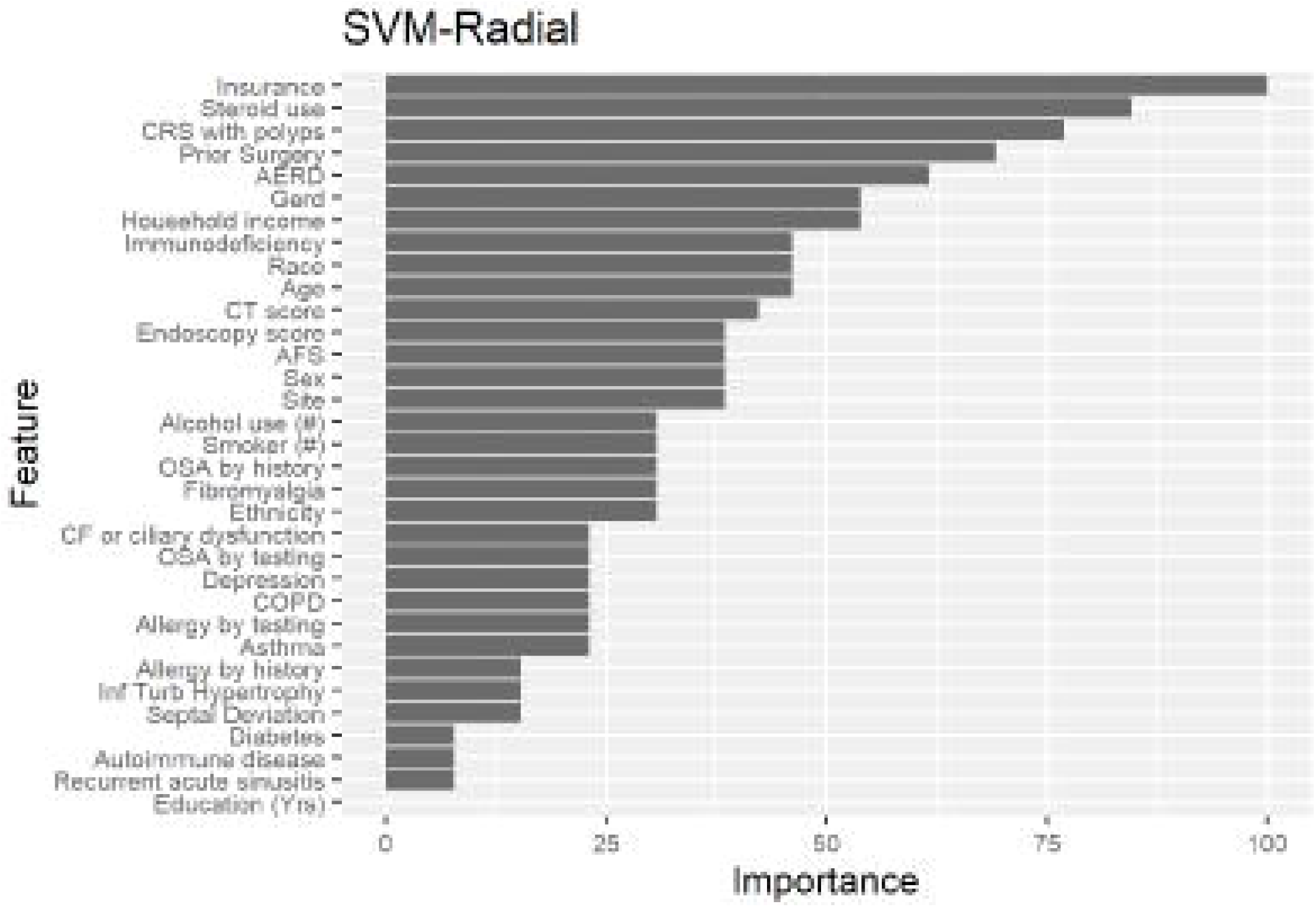
Variable importance display for most accurate classification model (Support Vector Machine with a radial basis kernel). Note the inclusion of many predictor variables – 32 are included with > 10% variable importance, suggesting potential interaction between predictors. *SVM-Radial = support vector machine with a radial basis kernel*.

**Figure 3.**
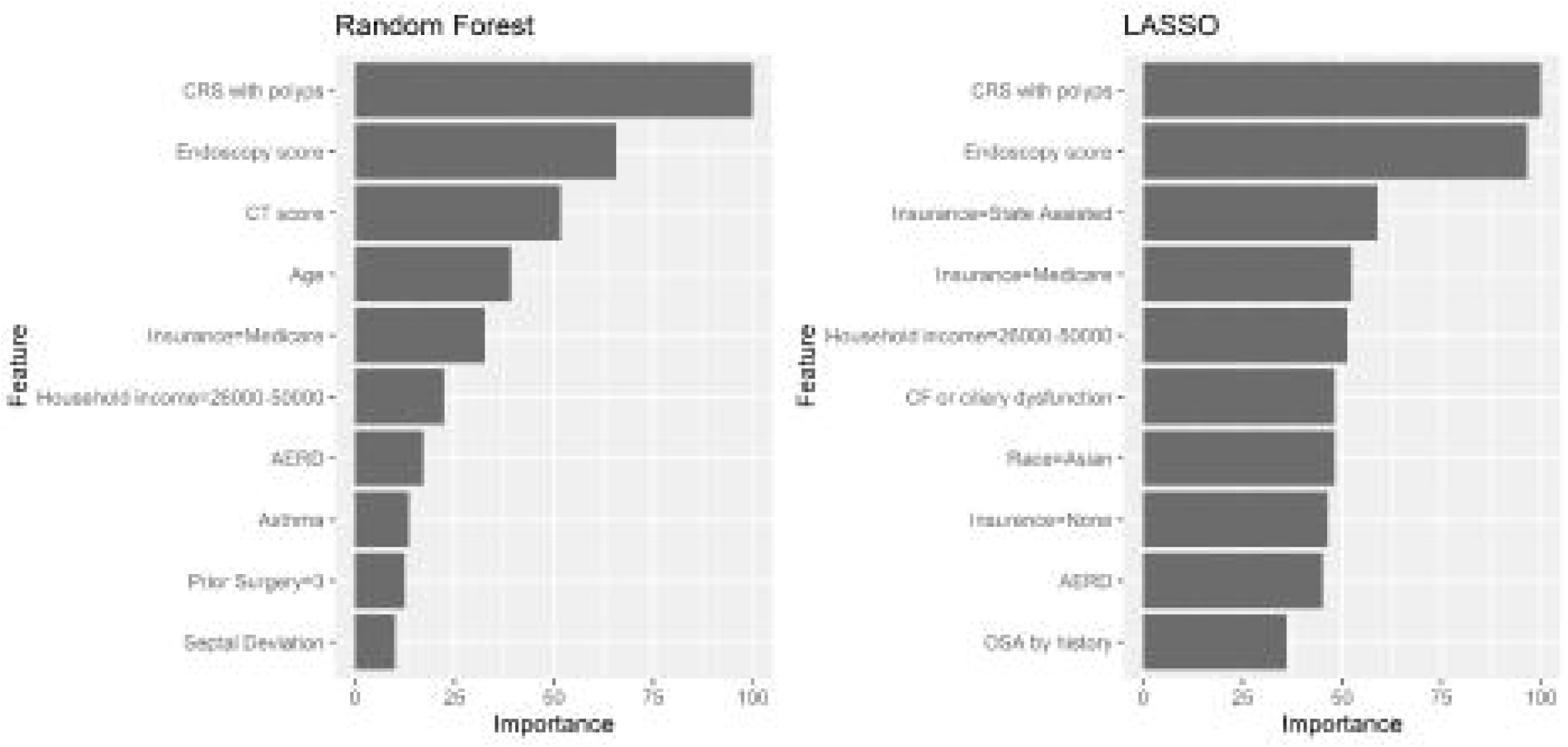
Top 10 Predictors and Variable importance for Random Forest and LASSO models. AERD = aspirin exacerbated respiratory disease; CF = cystic fibrosis.

Top predictors across the approaches include corticosteroid dependence, prior surgery, nasal polyps, AERD, age, sex, and smoking history. Inflammatory disease severity, as measured by CT and nasal endoscopy scores, were also strong predictors of CRS-OD.

Socioeconomic factors such as insurance status, household income, and race are among top predictors across the three models displayed. Age was a less important variable in all models. Comorbid medical conditions including asthma, diabetes, septal deviation, alcohol use, and sleep apnea, are also of moderate importance. Although the presence of nasal polyps, asthma, and AERD, are significantly associated with CRS-OD, the presence of allergic rhinitis by history or by skin testing appears to carry relatively minimal weight in the CRS-OD classifications.

## DISCUSSION

CRS is a heterogeneous process in terms of presentation, subtypes, natural history and response to treatments [2]. Current data from a multi-institutional CRS cohort studying olfactory loss in CRS with a sensitive olfactory test (Sniffin’ Sticks) demonstrates that nearly 70% of the CRS population has significant olfactory loss, with 20% exhibiting complete anosmia [7]. The pathophysiology of OD in CRS could result from both sensorineural and conductive effects of local tissue inflammation, on top of the myriad factors that can affect olfaction in the general population [1,46]. In previous studies, factors associated with worse baseline olfactory function in CRS patients included nasal polyposis, asthma, age, smoking, and eosinophilia [47]. Other works have shown CRS-OD response treatment in ∼40% of patients, with risk factors such as olfactory dysfunction severity, nasal polyps, female gender, high socioeconomic status and non-smoking associating with better quality of life results. In multivariate regression, only nasal polyps and degree of baseline olfactory dysfunction maintained statistical significance, highlighting the need for robust sample size and improved analytical methods to glean useful information from human study [10].

Here, we utilized ML-based data science approaches in addition to logistic regression in predictive modeling of CRS-OD classification. Top predictors across the approaches contain many variables associated with olfactory function in previous studies including nasal polyps, prior surgery, AERD, age, sex, corticosteroid dependence, and smoking history. Inflammatory disease severity, as measured by CT and nasal endoscopy scores, are also apparently important predictors of CRS-OD. Socioeconomic factors such as insurance status, household income, and race were among the important variables across the three top performing models. Since age was less important in all models, insurance status appears to be an independent predictor (i.e., Medicare insurance is not simply a proxy for advanced age). Comorbid medical conditions including asthma, diabetes, septal deviation, alcohol use, and sleep apnea, are also of moderate importance. Notably, these factors can be influenced by medical care and lifestyle modification, suggesting that other approaches to improve general health and well-being may be considered as part of a holistic approach to CRS-OD management. Although the presence of nasal polyps, asthma, and AERD, are significantly associated with CRS-OD here and in prior literature, the presence of allergic rhinitis by history or by skin testing appeared to carry relatively minimal weight in the CRS-OD classifications. This observation suggests that evaluation and treatment for allergic rhinitis may not necessarily be expected to benefit olfactory function in CRS.

We observed favorable prediction accuracy with most ML approaches compared to traditional regression modeling. ML is defined as the use of algorithms to break down data, learn from it, and then make a determination or prediction about some aspect. Whereas traditional statistical methods focus on modeling how system variables relate to one another and what statistical inference (e.g. significance in p-values) can be made, the goal of machine learning (ML) is not interpretation of individual components but prediction of future outcomes. In doing so, ML provides a novel approach to uncover previously unrecognized patterns among CRS-OD patients and thereby offers numerous advantages over regression analyses, which have been traditionally employed in studies of CRS disease severity and outcomes [48]. As more complex and multi-dimensional data are adding in “deep phenotyping” approaches to CRS, machine learning approaches may be well-suited to include the expansive amounts of added data (i.e., multi-’omics). Further external validation using appropriate cohorts will certainly be of value.

In this study, four different supervised ML algorithms were used map features of interest to the outcome or “label” of olfactory loss, and three outperformed traditional regression. In particular, the SVM model had improved accuracy that was driven largely by improved sensitivity. The classification accuracy may be attributed to the ability of these ML models to handle high-dimensional data in which more features exist than observations, and include non-linear and interaction effects. In contrast, traditional statistical modeling with a large number of potential features requires some form of dimension reduction or variable selection, and exploration of interactions and non-linear effects is challenging with more than a small number of predictors. As a result, we believe that ML and artificial intelligence data analytics are well-suited to prime research and eventually for the application of precision medicine in olfactory disorders, given the idiosyncrasies, nuances, and numerous possible predictor variables.

There are some limitations of the current study that should be addressed with regard to ML. Besides the four ML models considered in this study, many other ML methods exist and could be evaluated in future studies. Adequate enrollment by sex and race are common sampling issues in healthcare ML [49,50]. Details of this cohort have been extensively published and include even distributions by age and sex. Racial/ethnic minorities and low socioeconomic status groups are underrepresented perhaps as a function of geography and referral patterns of the enrollment sites, but also result from the general lack of accessible and affordable healthcare and racial disparities inherent within the U.S. healthcare system [51,52]. A related concern is that models created from our cohort data may “overfit” when new or unseen clinical data are applied. Certainly, these approaches should be further validated in independent studies, but nonetheless demonstrate the utility of data science approaches to uncover patterns within expansive human data in a noisy disease process. To incorporate a large enough patient dataset across three sites over several years of prospective data collection, psychophysical testing included classification of smell loss by either SIT or bSIT. Although the SIT may be more sensitive despite both of these tests being extensively validated in the literature, our prior analysis in a CRS cohort has demonstrated excellent diagnostic accuracy of the bSIT (AUC=0.873; 95% CI: 0.819, 0.927) [21]. Additionally, there may be important data inputs (i.e., molecular endotyping data) that are not yet clinically available. Serum markers, such as peripheral eosinophilia and serum IgE, have been proposed for CRS in the past but have generally been disappointing and do not appear to correlate with tissue levels [53,54]. Some of the clinical data input fields may associate phenotypes with likely molecular endotypes, but such overlap is not universal [5]. Future goals include testing whether additional accuracy is gained with the addition of molecular based biomarkers to the input data.

## CONCLUSION

Here we demonstrate the utility of novel predictive analytics approaches in the study of clinical olfactory disorders. Founded in data science principles rather than traditional statistics, ML models performed favorably compared to logistic regression. Consistently observed predictors across models include a weighted interest in socioeconomic status, and other potentially modifiable conditions such as asthma/AERD. Further studies will be important to reproduce and validate these findings, and might explore why certain novel predictor variables appear to be important. Such approaches may have value for both clinical counseling and guidance for future basic science and translational research.

## Data Availability

No datasets were generated during the current study. Due to the nature of the primary research study, participants did not agree for their data to be shared publicly, so these supporting data are not available.

## ACKNOWLEDGMENTS

The authors would like to thank Drs. Miranda Kroehl, PhD, John Rice, PhD, and Debashis Ghosh, PhD, for biostatistical expertise in discussions regarding validation of machine learning and prediction models. We would also like to thank Drs. Peter Hwang, MD, Rod Schlosser, MD, and Luke Rudmik, MD, for their contributions in original study enrollment and ongoing collaboration.

## TABLE AND FIGURE LEGEND

**Supplementary Figure 1.**
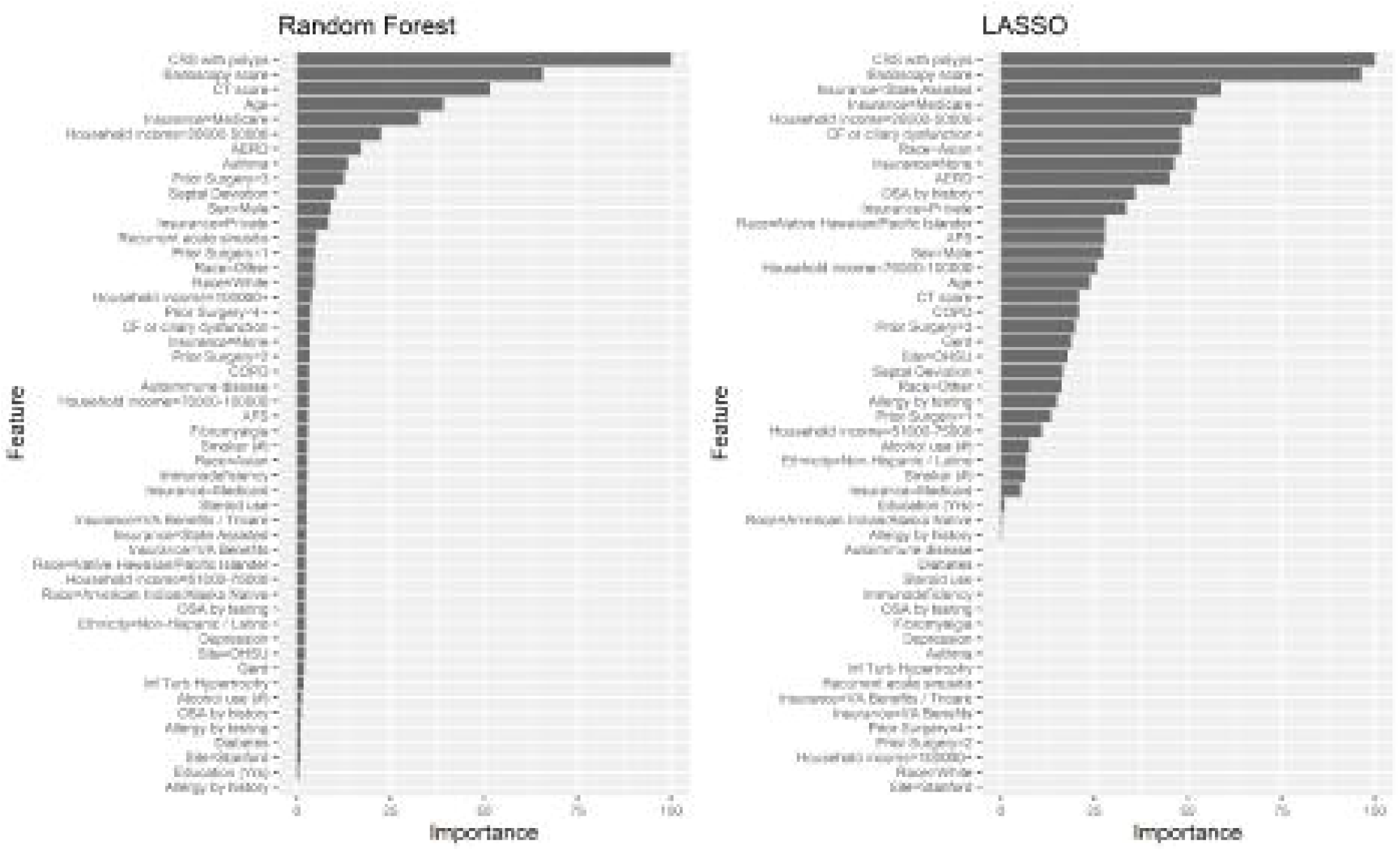
Complete list of predictors for Random Forest and LASSO models. Notably, the LASSO model incorporates significantly more features of interest than the Random Forest algorithm (26 features with >10% importance in the prediction algorithm).

## Notes

Financial Disclosures: This study was supported in part by a grant from the Ludeman Family Center for Women’s Health Research at the University of Colorado Anschutz Medical Campus (V.R.R.). V.R.R., J.C.M., Z.M.S., T.L.S., and S.S.S. are supported by grants for this investigation from the National Institute on Deafness and Other Communication Disorders (NIDCD) and the National Institute of Allergy and Infectious Diseases (NIAID) of the National Institutes of Health, Bethesda, MD., USA [R01 DC005805 (T.L.S. and Z.M.S.), K23 DC014747 (V.R.R.), 1P01AI145818-01 (S.S.S.)]. Public clinical trial registration (www.clinicaltrials.gov) ID# NCT01332136. Contents are the authors’ sole responsibility and do not necessarily represent official NIH views.

**Conflicts of Interest:** None related to this study

### Competing Interest Statement

The authors have declared no competing interest.

### Clinical Trial

NCT01332136

### Funding Statement

V.R.R., J.C.M., Z.M.S., T.L.S., and S.S.S. are supported by grants for this investigation from the National Institute on Deafness and Other Communication Disorders (NIDCD) and the National Institute of Allergy and Infectious Diseases (NIAID) of the National Institutes of Health, Bethesda, MD., USA [R01 DC005805 (T.L.S.), K23 DC014747 (V.R.R.), 1P01AI145818-01 (S.S.S.)].

### Author Declarations

COMIRB Protocol 19-2085

### Summary of Updates

Additional validation analyses performed, and revision of Figure 1

